# Prediction of immunotherapy response using deep learning of PET/CT images

**DOI:** 10.1101/2020.10.09.20209445

**Authors:** Wei Mu, Lei Jiang, Yu Shi, Ilke Tunali, Jhanelle E. Gray, Evangelia Katsoulakis, Jie Tian, Robert J. Gillies, Matthew B. Schabath

## Abstract

Currently only a fraction of patients with non-small cell lung cancer (NSCLC) experience durable clinical benefit (DCB) from immunotherapy, robust biomarkers to predict response prior to initiation of therapy are an emerging clinical need. PD-L1 expression status from immunohistochemistry is the only clinically approved biomarker, but a non-invasive complimentary approach that could be used when tissues are not available or when the IHC fails and can be assessed longitudinally would have important implications for clinical decision support. In this study, ^18^F-FDG-PET/CT images and clinical data were curated from 697 NSCLC patients from three institutions. Utilizing PET/CT images, a deeply-learned-score (DLS) was developed by training a small-residual-convolutional-network model to predict the PD-L1 expression status, which was further used to predict DCB, progression-free survival (PFS), and overall survival (OS) in both retrospective and prospective test cohorts of immunotherapy-treated patients with advanced stage NSCLC. This PD-L1 DLS significantly discriminated PD-L1 positive and negative patients (AUC≥0.82 in all cohorts). Further, higher PD-L1 DLS was significantly associated with higher probability of DCB, longer PFS, and longer OS. The DLS combined with clinical characteristics achieved C-indices of 0.86, 0.83 and 0.81 for DCB prediction, 0.73, 0.72 and 0.70 for PFS prediction, and 0.78, 0.72 and 0.75 for OS prediction in the retrospective, prospective and external cohorts, respectively. The DLS provides a non-invasive and promising approach to predict PD-L1 expression and to infer clinical outcomes for immunotherapy-treated NSCLC patients. Additionally, the multivariable models have the potential to guide individual pre-therapy decisions pending in larger prospective trials.

**Statement of Significance:** PD-L1 expression status by immunohistochemistry (IHC) is the only clinically-approved biomarker to trigger immunotherapy treatment decisions, but a non-invasive complimentary approach that could be used when tissues are not available or when the IHC fails and can be assessed longitudinally would have important implications for clinical decision support. Utilizing PET/CT images, we developed and tested a convolutional neural network model to predict PD-L1 expression status with high accuracy in cohorts from different institutions. And the generated signature may serve as a prognostic biomarker for immunotherapy response in patients with NSCLC, and outperforms the clinical characteristics.

## Introduction

The emergence of immune checkpoint inhibitors has revolutionized cancer treatment and improved long-term survival among some patients with advanced stage non-small cell lung cancer (NSCLC), but durable clinical benefit (DCB) has only been observed in 20-50% patients(1-4). Because of the complexity and heterogeneity in immunotherapy response and progression, robust and predictive biomarkers are urgently needed to identify patients who are unlikely to respond, and this is especially true for those patients that may experience rapid and lethal hyperprogressive disease(5).

At present, PD-L1 expression status by immunohistochemistry (IHC) is the only clinically-approved biomarker to trigger treatment decisions(3, 6), and early studies showed that PD-L1 positivity is associated with significantly higher objective response rate (ORR), longer progression-free survival (PFS) and longer overall survival (OS)(2, 7). However, measuring PD-L1 by IHC requires surgical or biopsied tumor specimens, which are collected through invasive procedures and associated with risk of morbidities. Additionally, IHC may not be possible due to poor quality tissue specimens or if tissues are unavailable. Additionally, the PD-L1 expression may change over the course of therapy and progression, which would require additional invasive sampling(8). Therefore, an alternative and complimentary non-invasive approach that could be used when tissues are not available or when the IHC fails and can be assessed longitudinally would have important implications for clinical decision support (9).

Quantitative image-based biomarkers (Radiomics) have many advantages over tissue-based biomarkers, like PD-L1, as they reflect underlying pathophysiology and tumor heterogeneity of the entire tumor and not just the portion of the tumor that is biopsied. Further, they can be rapidly calculated from standard-of-care medical images, such as ^18^F-FDG PET/CT, and can be captured longitudinally throughout the course of therapy to characterize response dynamics. Prior studies have shown that radiomic signatures based on shape, size, voxel intensity, and texture typically extracted from the intratumoral region are statistically associated with gene expression, tumor microenvironment, and treatment response in NSCLC(10-12). Additionally, it is becoming increasingly appreciated that the peritumoral region(13), encompassing the tumor-stroma interface, is important to capture and quantify, since this region contains immune infiltration and stromal inflammation. Intratumoral and peritumoral immune-cell infiltration is necessary for inducing an immunotherapy response. Immune infiltration is associated with expression of cell checkpoint markers including PD-L1(14), which is significantly correlated with metabolic rate, GLUT-1(15), pAKT(16), hypoxia, and acidosis(17). In light of the consolidated mutual interaction of metabolic and immune pathways, metabolic parameters SUVmax on FDG-PET was found statistically significantly associated with intra-tumor expression of immune-related markers (18). Recent studies have demonstrated several textural features from ^18^F-FDG PET/CT images can provide supplemental information to determine tumor PD-L1 expression (19, 20). Thus, ^18^F-FDG PET/CT radiomics could be particularly sensitive in predicting PD-L1 status due to its metabolic sensitivity in both intratumoral and peritumoral regions, which may identify patients who are most likely to benefit from immune checkpoint inhibitors.

To achieve this, we utilized artificial intelligence (AI) methods to develop and validate a deeply learned score (DLS) to predict PD-L1 expression status using pre-treatment ^18^F-FDG PET/CT images of a retrospective cohort accrued from Shanghai Pulmonary Hospital (SPH). To evaluate the performance of the PD-L1 status prediction model, two external test cohorts from H. Lee Moffitt Cancer Center & Research Institute (HLM) and James A. Haley Veterans’ Hospital (VA) are used. To determine its potential clinical utility for identifying patients most like to benefit from immunotherapy (IO), we tested the DLS to predict DCB, PFS and OS in two retrospective cohorts from HLM and VA and one prospective cohort from HLM (Details shown in **Fig. 1**).

**Fig 1.**
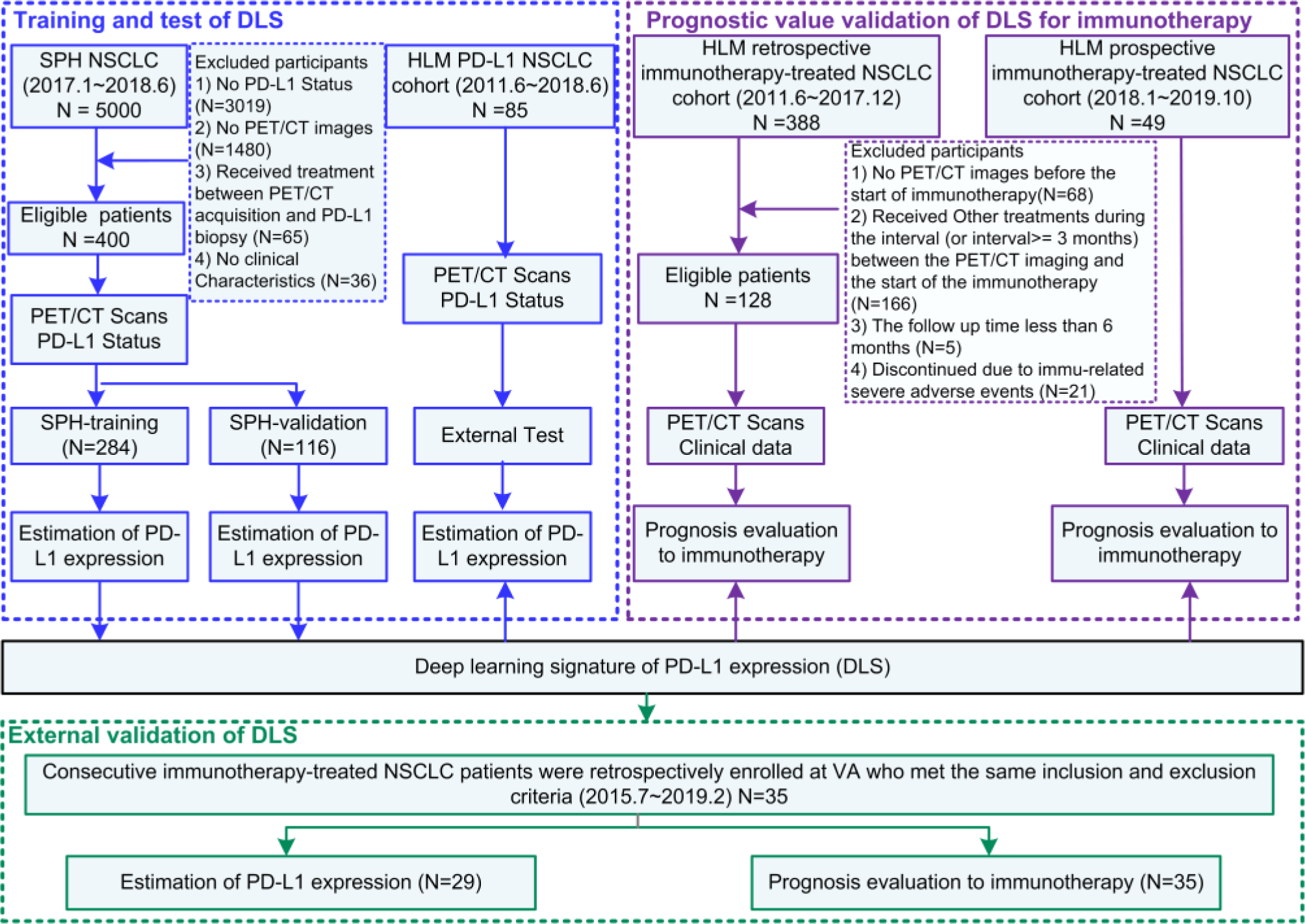
Study design and inclusion and exclusion diagram. The SPH data comprised PD-L1 expression data and the corresponding imaging data was used to train the deep learning signature. The HLM-PD-L1 data comprised PD-L1 expression data and the corresponding imaging data and was used for the test of the deep learning signature. The HLM retrospective and prospective data comprised patients included in anti-PD-1 and anti-PD-L1 immunotherapy were used for the investigation of the prognostic value of the deep learning signature. The external VA immunotherapy-treated data were used as the external validation for the estimation of PD-L1 expression and prognosis evaluation to immunotherapy.

## Results

### Patients characteristics

The clinical characteristics of the patients used to train and test the predictor for PD-L1 status are presented in **Table 1 (Supplemental Table S1** for external VA patients). The SPH-training, SPH-validation, and external HLM-PD-L1 test cohorts used to train, validate and test the SResCNN model had a prevalence of PD-L1 positivity by IHC of 29.93%, 30.17% and 54.12%, respectively. The external VA patients had a significant higher PD-L1 positivity of 82.76% (within the 29 patients who had IHC PD-L1 expression).

**Table 1.** Demographic and clinical characteristics of patients used to predict PD-L1 status.

| Characteristic | SPH Training Cohort (N=284) |  |  | SPH Validation Cohort (N=116) |  |  | HLM-PD-L1-test Cohort (N=85) |  |  |
| --- | --- | --- | --- | --- | --- | --- | --- | --- | --- |
|  | PD-L1 + | PD-L1 - | P | PD-L1 + | PD-L1 - | P | PD-L1 + | PD-L1 - | P |
| **Age(y)** |  |  | 0.41 |  |  | 0.46 |  |  | 0.40 |
| Mean±SD | 62.71±8.78 | 63.51±8.56 |  | 63.6±9.28 | 62.73±8.84 |  | 68.22±9.25 | 64.77±14.35 |  |
| **Sex, NO. (%)** |  |  | 0.035* |  |  | 0.062 |  |  | 0.082 |
| Male | 58 (68.24) | 108 (54.27) |  | 26 (74.29) | 44 (54.32) |  | 23 (50) | 27 (71.05) |  |
| Female | 27 (31.76) | 91 (45.73) |  | 9 (25.71) | 37 (45.68) |  | 23 (50) | 11 (28.95) |  |
| **TNM stage** |  |  | 0.12 |  |  | 0.42 |  |  | 0.25 |
| I | 43 (50.59) | 122 (61.31) |  | 17 (48.57) | 55 (67.9) |  | 0 (0) | 2 (5.26) |  |
| II | 22 (26.19) | 34 (17.08) |  | 9 (25.71) | 14 (17.28) |  | 2 (4.35) | 0 (0) |  |
| III | 11 (13.10) | 31 (15.58) |  | 9 (25.71) | 12 (14.81) |  | 2 (4.35) | 1 (2.63) |  |
| IV | 9 (10.71) | 12 (6.03) |  | 0 (0) | 0 (0) |  | 42 (91.3) | 35 (92.11) |  |
| **Histology (baseline), NO. (%)** |  |  | <.001* |  |  | 0.006* |  |  | 0.2 |
| ADC | 48 (56.47) | 156 (78.39) |  | 19 (54.29) | 65 (80.25) |  | 21 (42.86) | 26 (66.67) |  |
| SCC | 37 (43.53) | 43 (21.61) |  | 16 (45.71) | 16 (19.75) |  | 25 (57.14) | 13 (33.33) |  |
| **EGFR, NO. (%)** |  |  | 0.020 |  |  | 0.28 |  |  | 1.00 |
| Mutation | 24 (28.57) | 87 (43.72) |  | 9 (25.71) | 32 (39.51) |  | 2 (4.35) | 2 (5.13) |  |
| Wild | 55 (65.48) | 101 (50.75) |  | 23 (65.71) | 46 (56.79) |  | 37 (80.43) | 27 (69.23) |  |
| **ALK, NO. (%)** |  |  | 0.51 |  |  | 0.20 |  |  | 1.00 |
| Mutation | 1 (1.19) | 1 (0.50) |  | 2 (5.71) | 1 (1.23) |  | 1 (2.17) | 0 |  |
| Wild | 78 (92.86) | 187 (93.97) |  | 30 (85.71) | 77 (95.06) |  | 37 (80.43) | 31 (79.49) |  |
| **ROS1, NO. (%)** |  |  | 1.00 |  |  | NaN |  |  | NaN |
| Mutation | 0 | 2 (1.01) |  | 0 | 0 |  | 0 | 0 |  |
| Wild | 77 (91.67) | 179 (89.95) |  | 31 (88.57) | 75 (92.59) |  | 24 (52.17) | 19 (48.72) |  |
| **Smoking Status, NO. (%)** |  |  | <.001* |  |  | 0.025* |  |  | 0.12 |
| Never | 32 (37.65) | 118 (59.3) |  | 11 (31.43) | 45 (55.56) |  | 13 (41.94) | 33 (61.11) |  |
| Former | 53 (62.35) | 81 (40.7) |  | 24 (68.57) | 36 (44.44) |  | 18 (58.06) | 21 (38.89) |  |
| **SUVmax** |  |  | <.001* |  |  | 0.003* |  |  | 0.002* |
| Mean±SD | 12.32±5.99 | 8.42±4.83 |  | 12.14±5.88 | 8.61±5.49 |  | 14.26±7.8 | 10.14±9.64 |  |
| **Deep Learning Score** |  |  | <.001* |  |  | <.001* |  |  | <.001* |
| Median | 0.70 | 0.43 |  | 0.63 | 0.41 |  | 0.58 | 0.39 |  |
| (IQR) | (0.60,0.78) | (0.30,0.55) |  | (0.55,0.71) | (0.27,0.52) |  | (0.53,0.62) | (0.26,0.43) |  |
| **PD-L1 Positivity by IHC** |  |  |  |  |  |  |  |  |  |
| NO. (%) | 84 (29.93) | 199 (70.07) |  | 35 (30.17) | 81 (69.83) |  | 46 (54.12) | 39 (45.88) |  |

Note: \* means P value <0.05. The comparison of age and SUVmax between two groups was performed with Wilcoxon sign rank test, and the rest variables were compared with Fisher's exact test. IQR is short for interquartile range. The demographic and clinical characteristics of external VA patients were provided in Supplemental Table S2.

The clinical characteristics of the patients used to test the clinical utility of DLS are presented in **Table 2**. The retrospective HLM IO-treated cohort included 128 patients with a median PFS and OS of 7.43 and 21.77 months, respectively, and 53.91% of the patients had DCB. The prospective HLM IO-treated patients included 49 patients with a DCB rate of 65.31%, median PFS and OS of 7.93 and 17.00 months, respectively. For the external VA patients with a median PFS and OS of 8.13 and 13.10 months, 68.57% of the patients showed PD-L1 positive, and 54.29% of patients obtained DCB.

**Table 2.** Demographic and clinical characteristics for IO-treated patients.

| Characteristic | Retrospective HLM IO-treated patients (N=128) |  |  |  | Prospective HLM IO-treated patients (N=49) |  |  |  |
| --- | --- | --- | --- | --- | --- | --- | --- | --- |
|  | All | Deep Learning Score |  | P | All | Deep Learning Score |  | P |
|  |  | High (N=44) | Low (N=84) |  |  | High (N=32) | Low (N=17) |  |
| **Age(y)** |  |  |  | 0.18 |  |  |  | 0.20 |
| Mean ± SD | 65.48±13.24 | 67.11±13.75 | 64.63±12.96 |  | 66.8±10.04 | 65.31±9.38 | 70.00±10.42 |  |
| **BMI** |  |  |  | 0.17 |  |  |  | 0.48 |
| Mean ± SD | 26.14±5.08 | 25.40±5.39 | 26.53±4.90 |  | 26.06±5.02 | 25.97±5.47 | 26.1±4.09 |  |
| **Sex, NO. (%)** |  |  |  | 0.46 |  |  |  | 0.37 |
| Male | 81 (63.28) | 25 (56.82) | 56 (66.67) |  | 25 (51.02) | 18 (56.25) | 7 (41.18) |  |
| Female | 47 (36.72) | 19 (43.18) | 28 (33.33) |  | 24 (48.98) | 14 (43.75) | 10 (58.82) |  |
| **TNM stage** |  |  |  | 0.10 |  |  |  | 0.65 |
| III | 25 (19.53) | 11 (25.00) | 14 (16.67) |  | 6 (12.24) | 5 (15.63) | 1 (5.88) |  |
| IV | 103 (80.47) | 33 (75.00) | 70 (83.33) |  | 43 (87.76) | 27 (84.38) | 16 (94.12) |  |
| **Histology (baseline), NO. (%)** |  |  |  | 0.089 |  |  |  | 0.37 |
| ADC | 80 (62.50) | 23 (52.27) | 57 (67.86) |  | 28 (57.14) | 20 (62.5) | 8 (47.06) |  |
| SCC | 48 (347.50) | 21 (47.73) | 27 (32.14) |  | 21 (42.86) | 12 (37.5) | 9 (52.94) |  |
| **EGFR, NO. (%)** |  |  |  | 0.71 |  |  |  | 0.53 |
| Mutation | 8 (6.25) | 3 (6.98) | 5 (5.88) |  | 2 (4.08) | 2 (6.25) | 0 |  |
| Wild | 85 (66.41) | 27 (62.79) | 58 (68.24) |  | 37 (75.51) | 23 (71.88) | 14 (82.35) |  |
| **ALK, NO. (%)** |  |  |  | 1.00 |  |  |  | NaN |
| Mutation | 2 (1.56) | 0 | 2 (2.35) |  | 0 | 0 | 0 |  |
| Wild | 89 (69.53) | 28 (65.12) | 61 (71.76) |  | 39 (79.59) | 24 (75.00) | 15 (88.24) |  |
| **ROS1, NO. (%)** |  |  |  | NaN |  |  |  | NaN |
| Mutation | 0 | 0 | 0 |  | 0 | 0 | 0 |  |
| Wild | 35 (27.34) | 7 (16.28) | 28 (32.94) |  | 33 (67.35) | 20 (62.50) | 13 (76.47) |  |
| **Smoke, NO. (%)** |  |  |  | 0.70 |  |  |  | 0.74 |
| Never | 49 (38.28) | 18 (40.91) | 31 (36.9) |  | 14 (28.57) | 10 (31.25) | 4 (23.53) |  |
| Former | 79 (61.72) | 26 (59.09) | 53 (63.1) |  | 35 (71.43) | 22 (68.75) | 13 (76.47) |  |
| **ECOG Scale, NO. (%)** |  |  |  | 0.15 |  |  |  | 0.72 |
| 0 | 29 (22.66) | 6 (13.64) | 23 (27.38) |  | 10 (16.33) | 6 (18.75) | 4 (23.53) |  |
| 1 | 91 (71.09) | 36 (81.82) | 55 (65.48) |  | 38 (81.63) | 26 (81.25) | 14 (82.35) |  |
| ≥2 | 8 (6.25) | 2 (4.55) | 6 (7.14) |  | 1 (2.04) | 1 (3.13) | 0 (0) |  |
| **SUVmax** |  |  |  | 0.005* |  |  |  | 0.15 |
| Mean ± SD | 11.82±6.98 | 13.53±5.41 | 10.93±7.55 |  | 14.59±9.53 | 15.21±7.55 | 13.44±12.65 |  |
| **Clinical Benefit, NO. (%)** |  |  |  | <.001* |  |  |  | 0.004* |
| DCB | 69 (53.91) | 33 (75.00) | 36 (42.86) |  | 32 (65.31) | 26 (81.25) | 6 (35.29) |  |
| NDB | 59 (46.09) | 11 (25.00) | 48 (57.14) |  | 17 (34.69) | 6 (19.75) | 11 (64.71) |  |
| **Progression free Survival** |  |  |  | <.001* |  |  |  | 0.011* |
| Median | 7.43 | 15.80 | 5.50 |  | 7.93 | 17.00 | 4.00 |  |
| (95%CI) | (6.39,8.47) | (9.67,21.93) | (2.87,8.13) |  | (4.67,11.20) | (4.25,29.75) | (2.39,5.61) |  |
| **Overall Survival** |  |  |  | 0.024* |  |  |  | <0.001* |
| Median | 21.77 | 27.60 (NR) | 19.77 |  | 17.00 (NR) | NR | 11.23 |  |
| (95%CI) | (13.50,30.03) |  | (13.58,25.96) |  |  |  | (6.60-15.86) |  |
| **Deep Learning Score** |  |  |  | <.001* |  |  |  | <.001* |
| Median | 0.48 | 0.63 | 0.42 |  | 0.55 | 0.59 | 0.33 |  |
| (IQR) | (0.01,0.93) | (0.54,0.93) | (0.01,0.54) |  | (0.14,0.86) | (0.54,0.86) | (0.14,0.52) |  |

Notes: \* indicates a P value <0.05. The comparison of age, BMI and SUVmax between two groups was performed with Wilcoxon sign rank test, PFS and OS were compared with log-rank test, and the rest variables were compared with Fisher's exact test. IQR: interquartile range. NR: Not reached

### Performance of DLS in predicting PD-L1 status

The DLS exhibited statistically significant differences between the PD-L1-positive and PD-L1-negative tumors in all three cohorts (*p<*0.001), and four examples are shown in **Fig 2**. To discriminate PD-L1 positive from negative expression, the DLS yielded AUCs of 0.89 (95%CI:0.84-0.94; *p<*0.001) and 0.84 (95%CI:0.76-0.92; *p<*0.001), and 0.82 (95%CI:0.74-0.89; *p<*0.001) in the SPH training and validation, and HLM-PD-L1 test cohort, respectively (**Fig 3** and **Supplemental Table S2**). For the external VA patients, the DLS also generated a high AUC of 0.84 (95%CI:0.69-0.99; *p=0*.*018*). As another meaningful quantitative index associated with PD-L1 expression validated in other studies(21), SUV_max_ showed poorer performance to discriminate between PD-L1 positive and negative expression with AUCs of 0.69 (95%CI:0.62-0.75; *p*<0.001), 0.68 (95%CI:0.57-0.78; *p*<0.001), 0.69 (95%CI:0.58-0.81; *p*<0.001), and 0.48 (95%CI:0.2-0.78; p=0.86) in the SPH-training, SPH-validation, external HLM-PD-L1-test and VA cohorts, respectively.

**Fig 2.**
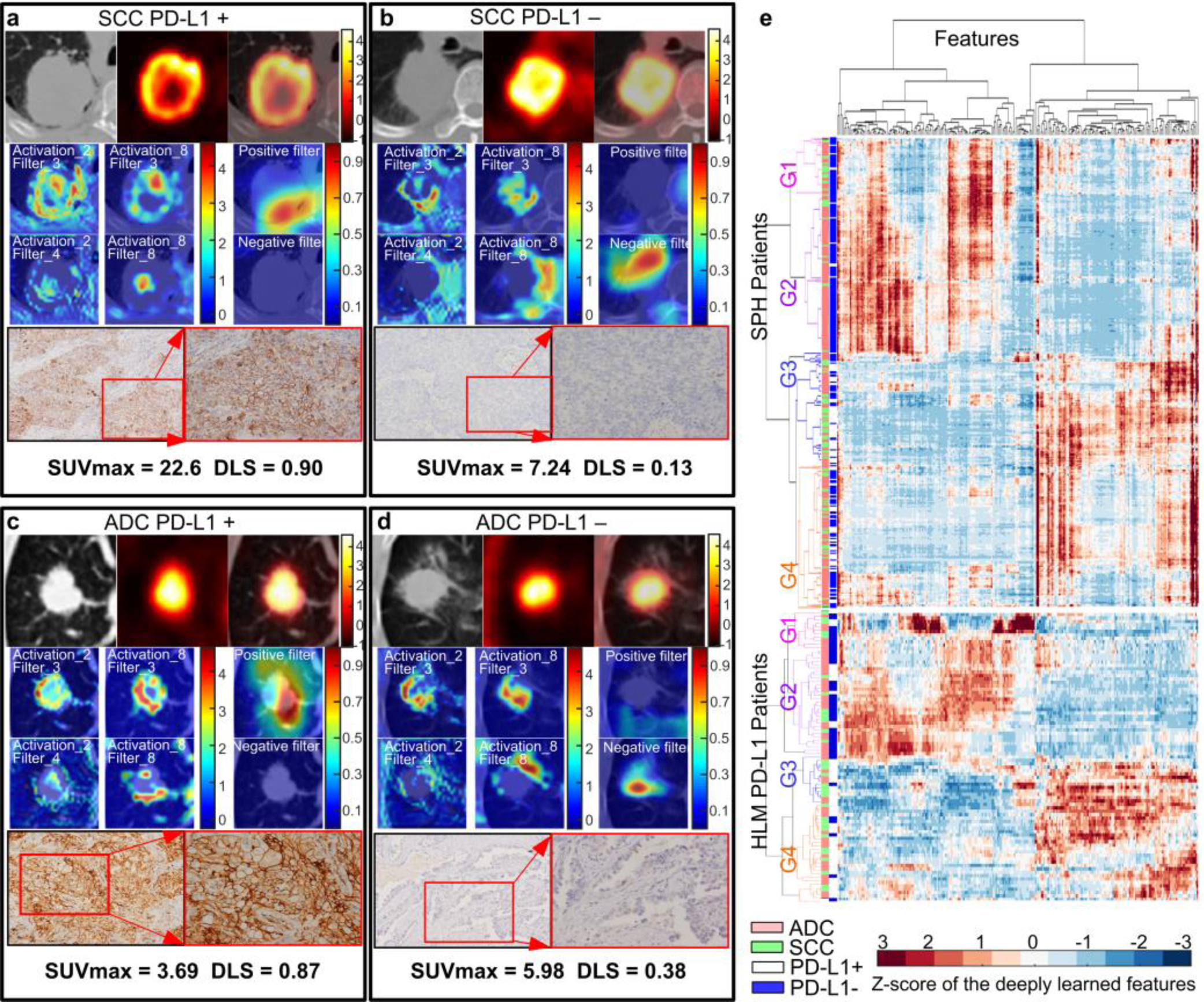
NSCLC histology subtypes and PD-L1 expression. Squamous cell carcinoma (SCC) patients with positive PD-L1 expression (a) and negative PD-L1 expression (b). Adenocarcinoma (ADC) patients with positive PD-L1 expression (c) and negative PD-L1 expression (d), respectively. For (a)-(d), the first line of (a)-(d) are the CT, PET and fusion images, the first and second columns of the second and third line shows the response of the fourth ResBlock, which shows the self-learned important areas in expressing PD-L1 status (peritumoral and necrosis regions), the third column of the second and third line shows the response of the negative filter and the positive filter in the PD-L1 positive-negative tumors (the CT images were overlapped to reveal the location of the response), the last line shows the pathological examination of the resected mass demonstrating PD-L1 expression (left, ×100; right, ×200). (e) The heatmap generated with unsupervised hierarchical clustering of all the SPH patients and HLM-PD-L1 patients on the horizontal axis and deeply learned features expression (i.e. the output of the last activation filters, N=256) on the vertical axis. There were four distinct subgroups obtained. Groups G1 and G2 (including more PD-L1-patients) had similar feature expression, which is opposite to the feature expression of G3 and G4 (including more PD-L1+ patients). Furthermore, some features of G1 and G2 (or G3 and G4) are different. G1 and G3 had more SCC patients, while G2 and G4 had more ADC patients. The χ2 test showed the significant association of the 4 kind of deep learning expression patterns with PD-L1 expression (SPH patients: *p<*0.001, HLM patients: *p<*0.001), and different histology (SPH patients: *p<*0.001, HLM patients: *p=*0.061). The similar patterns of the external HLM-PD-L1 cohorts further showed the stability of the deep learning features.

**Fig 3.**
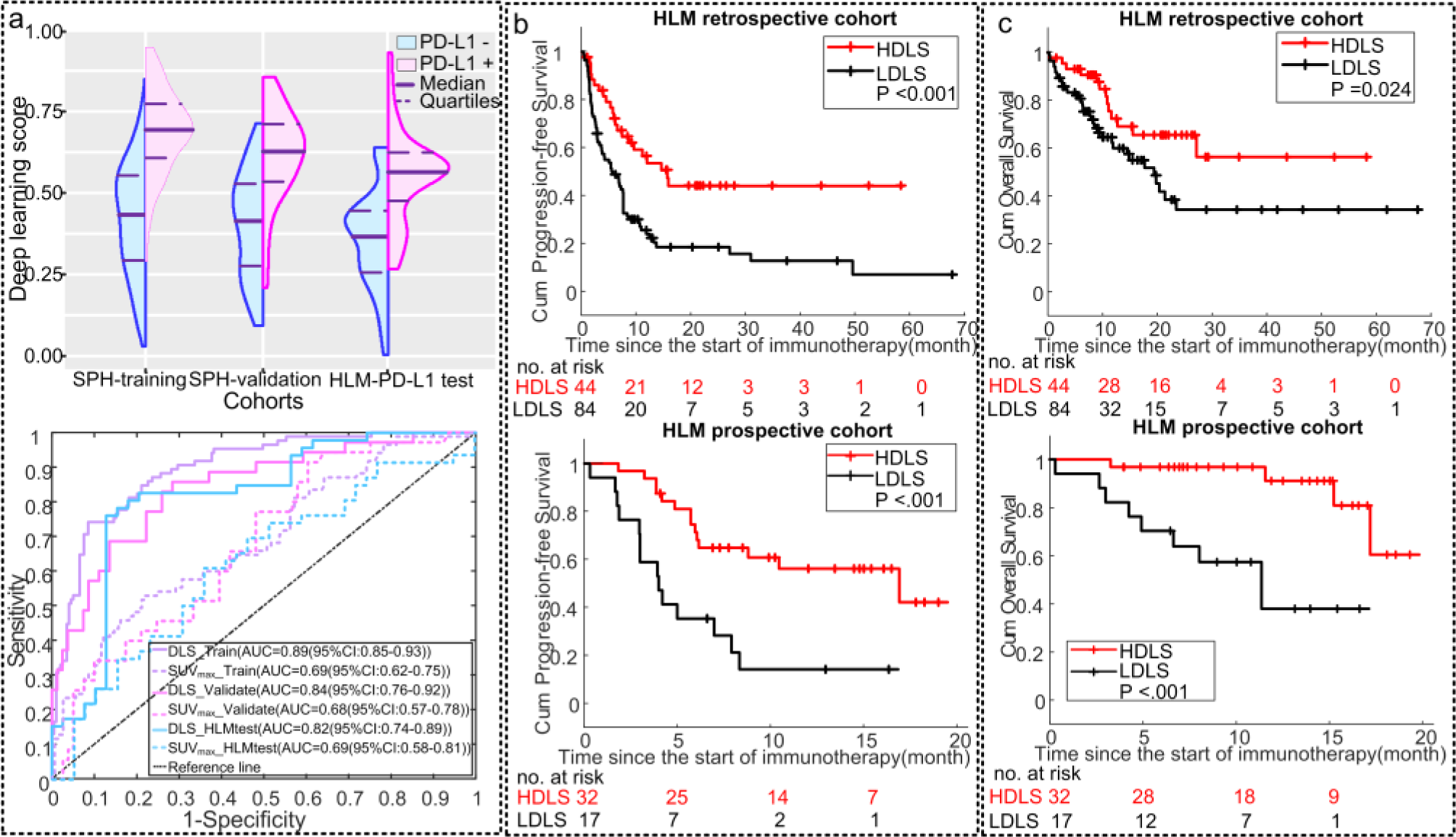
Performance of the DLS in the different cohorts. (a) The distribution of DLS between PD-L1 positive (+) and negative (-) groups in SPH-training, SPH-validation and external HLM-PD-L1-test cohorts, and the ROC curves of DLS and SUVmax in SPH-training, SPH-validation and external HLM-PD-L1-test cohorts. (b) The PFS relative to the DLS (high versus low defined by 0.54, which was the median value of the SPH training) in the retrospective and prospective HLM IO-treated patients. (c) The OS relative to the DLS (high versus low defined by 0.54, which was the median value of the SPH training) in the retrospective and prospective HLM IO-treated patients.

Since histology was found to be significantly associated with PD-L1 expression (*p*<0.01), a stratified analysis was conducted to assess the SResCNN model in predicting PD-L1 status by histology (**Supplemental Table S2**). The results from these analyses indicated this model also performed well in both adenocarcinoma (ADC) and squamous cell carcinoma (SCC) lung cancers.

Regarding the stability of the DLS, though accurate segmentations were not needed, radiologists had to delineate a rough ROI that contained the tumors and some surrounding tissue. To investigate the effect of the minor differences between the different radiologists in selecting the rough ROIs, the ROIs of the SPH validation patients (n=116 cases) were generated by two radiologists, and two DLSs were obtained accordingly. The intraclass correlation coefficient (ICC) of these two DLSs was 0.85 (95%CI:0.80-0.90, *p*<0.001), and showed no significant (p=0.85, Z-test) difference in the AUCs between different radiologists (0.84(95%CI:0.76-0.92) vs 0.83(95%CI:0.75-0.91)). Besides, there were no significant differences in the distribution of the DLS among the five cohorts (*p=*0.25, **Supplemental Fig S1**).

### Correlation between DLS and metadata and molecular biology

The DLS was also positively correlated with the original PD-L1 TPS in both SPH (Spearman’s rho=0.60, *p*<0.001) and HLM-PD-L1-test (Spearman’s rho = 0.59, *p*<0.001) cohorts, which was significantly higher compared to the correlation between the SUV max and the TPS with rho of 0.30 (*p*<0.001) and 0.33 (*p*=0.02), respectively. Using ANOVA, the DLS was significantly different between groups with PD-L1 TPS <1%, 1-49% and ≥50% (SPH cohort: P<0.001; HLM-PD-L1-test cohort: *p*<0.001). The least squares difference (LSD) *post hoc* analysis showed significantly higher values of DLS in the patients with PD-L1 TPS≥50% than TPS 1-49% group (LSD: SPH cohort: *p*<0.017; HLM-PD-L1-test cohort: *p*<0.026) and TPS<1% group (LSD: SPH cohort: *p*<0.001; HLM-PD-L1-test cohort: *p*<0.001) (Details shown in **Supplemental Fig S2**). As such the increased PD-L1 TPS scores correlated to the DLS.

Additionally, the DLS was positively correlated with SUVmax (rho=0.36, *p*<0.001), squamous cell carcinoma (rho=0.23, *p*<0.001), male sex (rho=0.17, *p*<0.001), smoking status (rho=0.17, *p*<0.001), and negatively correlated with EGFR status (rho=-0.17, *p*<0.001) for the whole SPH cohort. In the HLM-PD-L1 test cohort, the only positive correlation was with SUVmax (rho=0.35, *p* <0.001), and negative with EGFR status (rho=-0.24, *p*=0.49). Further, multivariable linear regression (adjusted r^2^=0.15, F=15.31, *p*<0.001) showed that only SUVmax (coefficient=0.32, *p*=0.005) was independently associated with DLS. Only 15% of DLS variability could be explained by this parameter indicating that DLS originated mainly from other image information.

For all the SPH patients with necrosis regions, a significant correlation was observed between the necrosis-to-global volume ratio (NVR) of the PET images and DLS with Spearman’s rho of 0.61 (*p*<0.001). Further, univariable linear regression (adjusted r^2^=0.22, F=8.93, *p*=0.005) showed that the necrosis (coefficient=0.47, *p*=0.005) was independently associated with DLS and could explain 22% of DLS variability. Therefore, the necrosis potentially played an important role in predicting PD-L1 status.

Finally, the DLS was not correlated with tumor volume (*p=*0.10 for whole SPH cohort, *p=*0.66 for HLM-PD-L1-test cohort), which shows that the DLS is not affected by the T stage of the primary tumor.

### Performance of DLS in IO-treatment response and patient outcomes prediction

The DLS in the patients experiencing DCB was significantly higher compared to those who did not in both the HLM retrospective (0.53 vs. 0.43, p<0.001) and prospective (0.57 vs. 0.45, *p*=0.014) cohorts. The AUCs of the DLS to identify the DCB patients were 0.70 (95%CI:0.63-0.77,*p*<0.001) and 0.72 (95%CI:0.62-0.84, *p*=0.014) in the retrospective and prospective patients, respectively.

For the retrospective patients, the PFS and OS were significantly longer among patients with high DLS (≥0.54) versus patients with low DLS (PFS: hazard ratio[HR]:0.42, 95%CI:0.26-0.69, *p*=0.001; OS: HR:0.49, 95%CI:0.26-0.92, *p*=0.028; **Fig 3**). Among patients with high DLS, the median PFS and OS were 15.80 months and 27.60 months, compared to 5.50 months and 19.77 months for patients with low DLS (PFS:*p*<0.001; OS:*p*=0.024). Similar results were also observed in the prospective patients with the HRs of 0.28 (95%CI:0.13-0.60, *p*=0.001) and 0.12 (95%CI:0.032-0.46, *p*=0.002) DLS for PFS and OS respectively (**Fig 3**). High DLS patients had a longer median PFS of 17.00 months compared to 4.0 months in the low DLS patients (*p*<0.001,**Fig 3**). Median time to an OS event was not reached in the high DLS group and was 11.2 months in the low DLS group (*p*<0.001, **Fig 3**). The external VA test patients further validate the prognostic value of DLS with HRs of 0.36 (95%CI:0.13-0.99, *p*=0.049) and 0.30 (95%CI:0.09-0.99, *p*=0.049) for PFS (9.00 vs 2.37 months, p=0.040) and OS(15.53 vs 4.93 months,*p*=0.037), respectively.

### Multivariable analysis for clinical outcomes prediction

Univariable logistic and Cox regression analyses of the clinical characteristics (**Supplemental Tables S3-S6**) and gene mutation showed that none of three gene mutations were associated with clinical outcome, that patients with lower ECOG Status and adenocarcinoma showed significantly longer OS and PFS. Stratified analyses by histology and ECOG Performance Status were thus performed to investigate the ability of DLS to predict outcomes in these subgroups. Among patients with ADC, the DCB rates were 91.3% and 100% in patients with higher DLS versus 50.88% and 62.5% in patients with lower DLS in both retrospective and prospective cohorts (p<0.001), respectively (**Fig 4**). Among SCC patients, though the DCB rates were lower compared to ADC, the patients with higher DLS still had a significantly higher DCB rates in both retro- and prospective cohorts. Congruously, the PFS and OS of high DLS group were also longer than the low DLS group in both ADC and SCC subgroups (**Supplemental Table S7**). The results of the stratified analysis based on ECOG status (**Supplemental Table S8**) also showed that low DLS was still associated with poor outcomes among patients with high ECOG status (≥1).

**Fig 4.**
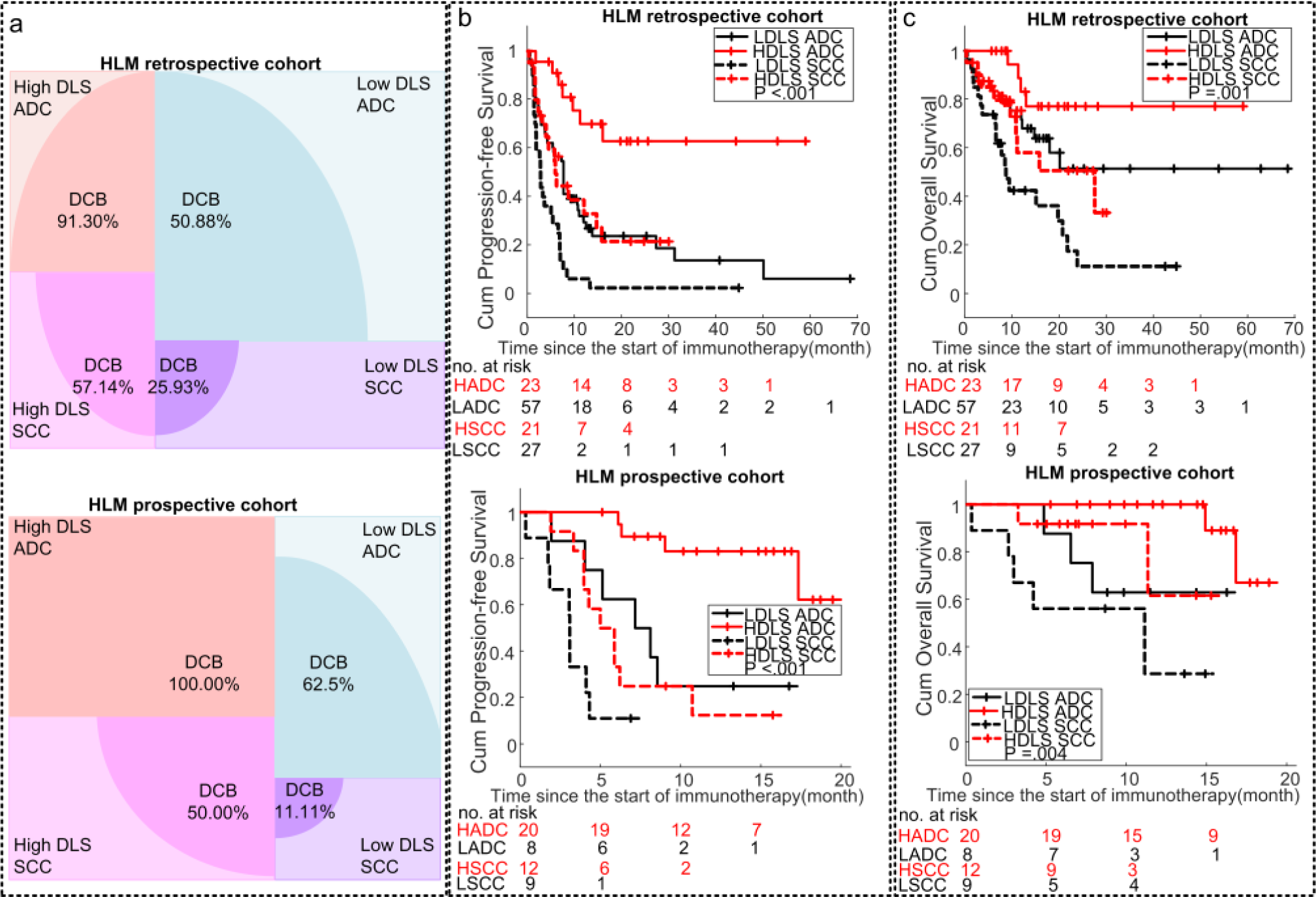
Stratification analysis of the performance of the DLS in prognosis prediction. (a) The DCB rates of the different subgroups of the HLM retrospective and prospective IO-treated patients. (b) The PFS relative to the DLS and histology in the HLM retrospective and prospective IO-treated patients. (c) The OS relative to the DLS and histology in the HLM retrospective and prospective IO-treated patients. Note: HADC is short for HDLS ADC, meaning ADC patients with high DLS, LADC is short for LDLS ADC, meaning ADC patients with Low DLS, HSCC is short for HDLS SCC, meaning SCC patients with High DLS, and LSCC is short for LDLS SCC, meaning SCC patients with Low DLS, the high DLS versus low DLS defined by 0.54, which was the median value of the SPH training.

Multivariable logistic regression and Cox proportional hazards regression analyses were conducted to adjust for potential confounding. Models including DLS, histology, and ECOG status, demonstrated high performance statistics (**Supplemental Table S9** and **Supplemental Fig S3**) with C-indices of 0.86 (95%CI:0.80-0.92, *p*<0.001) and 0.83 (95%CI:0.73-0.93, *p*<0.001) in DCB prediction, 0.73 (95%CI:0.69-0.78, *p*<0.001) and 0.72 (95%CI:0.64-0.82, *p*<0.001) in PFS prediction, 0.78 (95%CI:0.73-0.84, *p*<0.001) and 0.72 (95%CI:0.53-0.91, *p*<0.001) in OS prediction for the retrospective and prospective cohorts, respectively, showing better performance than clinical characteristics based models (including ECOG and histology) (*p*≤0.05, **Supplemental Table S10**). These models also demonstrated high performance statistics with C-indices of 0.81 (95%CI:0.70-0.93, *p*<0.001), 0.70 (95%CI:0.59-0.80, *p*<0.001), and 0.75 (95%CI:0.65-0.84, *p*<0.001) in DCB, PFS and OS prediction,respectively in the external VA patients.

## Discussion

Though PD-L1 expression status based on IHC is currently used as a clinical decision-making tool to support the use of checkpoint inhibitors in NSCLC patients, PD-L1 testing has inherent analytic and predictive limitations. As such, there is a pressing need to identify robust and reproducible biomarkers that are highly predictive of immunotherapy treatment response that may complement PD-L1 IHC. In this study we developed a deep learning model using PET/CT images to predict PD-L1 expression and found that the deeply learned score (DLS) could discriminate between positive and negative expression yielding an AUC of 0.89 in the training cohort and AUCs of 0.84 and 0.82 in two independent test cohorts. When the DLS was combined with clinical covariates and tested in two cohorts for clinical utility by identifying patients most like to benefit to immunotherapy, we found high C-indices of 0.83-0.86 for predicting DCB, but somewhat attenuated C-indices of 0.72-0.78 for the DLS to predict PFS and OS. These models also demonstrated good performance in the external VA cohort.

While others have demonstrated the utility of radiomics as a non-invasive approach to predict PD-L1 expression(11, 12) or predict lung cancer immunotherapy treatment response(22-26), the current work is the first to develop a PD-L1 radiomic signature and then to use this for response prediction. With respect to PD-L1 expression prediction, Patil et al. (11) utilized CT images from 166 early stage NSCLC patients from a single institution to develop and validate a machine learned predictor of PD-L1 status with an AUC of 0.73. Jiang et al.(12) utilized PET/CT radiomics of 399 NSCLC patients from a single institute to generate a classifier model with an AUC of 0.86. For the studies utilizing radiomics to predict immunotherapy treatment response, Tunali et al. built parsimonious classifier models with pre-treatment CT radiomic features combined with clinical covariates to predict hype-progression and progressive disease phenotypes with AUCs of 0.80-0.87(22), which were also highly correlated with PFS and OS of immunotherapy(23). Trebeschi et al.(24) developed a CT-based radiomic signature that significantly discriminated progressive disease from stable and responsive disease (AUC=0.83) among NSCLC patients treated with immunotherapy. He et al developed and tested CT-based Tumor mutational burden (TMB) predictor with 327 patients, which expresses prognostic value in PFS and OS prediction of immunotherapy in patients with advanced NSCLC(26). Notably, these prior studies were mostly limited to CT and required explicit tumor segmentation. By contrast, our study did not require accurate tumor segmentation, was conducted using rigorous training and validation in multiple cohorts from three institutions, and represented the single largest radiomic study population of NSCLC patients to date treated with immunotherapy to predict PD-L1 status and subsequent treatment response using ^18^F-FDG PET/CT. Further, these studies were trained to only predict outcome without regard to PD-L1 status; thus, not accommodating the only accepted biomarker to direct treatment with immune checkpoint blockade. We acknowledge that we observed a small attenuation in performance to predict PFS and OS compared to previous published radiomics model that was trained on clinical outcome without going through the process of predicting PD-L1(27); however, the current model has implications for clinical decision support because it can provide diagnostic information when tissue samples are insufficient, not available, or if an immediate PD-L1 score is needed.

Regarding underlying biology, one of the high-response areas of the middle layer of the SResCNN model recognized the necrotic region (activation_8_filter_8 in **Fig 2a-b**) through the visualization(28), suggesting that some final discriminant deeply learned features originate from necrotic regions. The quantification of the correlation between necrosis and DLS (Spearman’s rho: 0.61, p<0.001, univariable linear regression: coefficient=0.47, p=0.005) further potentially validated the important role of necrosis in predicting PD-L1 status. These results were also consistent with Jreige’s result that the metabolic-to-morphological volume was a predictive biomarker to predict PD-L1 expression(29). This could be explained with the presence of hypoxia, which can lead to necrotic cell death(30) and upregulate PD-L1 via hypoxia-inducible factor (HIF)-1α(31).

Additionally, the visualization of the SResCNN model (activation_8_filter_8 in **Fig 2c-d**) showed some final discriminant deeply learned features also originated from peritumoral region, and the high-response area of the positive/negative filter (**Fig 2c-d**) reconstructed by Grad-CAM also locate the peritumoral regions important regions, which suggests the peritumoral region may have played an important role in predicting PD-L1 expression. This is supported by prior work that higher levels of PD-L1+ staining in cells of peritumoral areas(32). These findings revealed an advantage of deeply learned models, which can agnostically capture features from the tumor and peritumoral microenvironments.

Furthermore, the response strength of the positive or negative filter varied with the histology for the same group of PD-L1 expression (**Fig 2a-d**), and the 4 main distinct subgroups of deep learned features indicated by unsupervised hierarchical clustering analysis were also significantly correlated with histology (**Fig 2e**). Within PD-L1 positive patients or PD-L1 negative patients, there was significant difference of DLS between different histologies (PL-L1 +: *p*=0.026, PD-L1 -: *p*=0.007), and the high DLS is significantly correlated with squamous cell carcinoma (PL-L1 +: Spearman’s rho =0.22, *p*=0.018, PD-L1 -: Spearman’s rho =0.16, *p*=0.006). The different feature expression of ADC and SCC tumors by PD-L1 status is consistent with the genomic differences between ADC and SCC revealed by large-scale sequencing studies(33). Additionally, better outcomes for ADC (Fig 4b-c) were observed which may be possibly due to a significantly higher tumor-infiltrating lymphocyte population estimated using immune cell signatures compared to SCC(34).

We also observed that hyper-image constructed with different modalities could significantly improve the accuracy of PD-L1 expression modelling. By training similar SResCNN models only using PET or CT images, the resulting DLSs (named DLS_PET and DLS_CT) achieved AUCs of 0.81 and 0.78 in the training cohort, 0.73 and 0.70 in the validation cohort, which was significantly worse (p<0.001) than those generated using the hyper-images. This may be attributed to the important regions (represented by necrosis and the peritumoral regions) used for the accurate prediction of PD-L1 expression could be better and easier localized by utilizing both metabolic and anatomical information as reflected by PET and CT images, respectively.

We do acknowledge some limitations of this study. First, the PD-L1 prediction training data are still limited to a single institution and EGFR mutations were highly prevalent in the Asian patient population at 40% compared to only 7% in whites, but there are no significant association between different ethnicities and PD-L1 expression(35). Additionally, the DLS could obtain high AUCs of 0.90, 0.87 and 1.00 in patients with mutated EGFR and also high AUCs of 0.88, 0.82, and 0.80 in the patient with wild type of EGFR in the SPH-training, SPH-validation and HLM-PD-L1-test cohorts, respectively. Therefore, the difference of ethnicity and EGFR positive patients’ percentage between the two institutional cohorts would not cause significant effect on PD-L1 expression prediction. Second, compared to other PD-L1 level detection methods, such as enzyme-linked immunosorbent assay (ELISA)(36), immunofluorescence (IF)(37), and flow cytometry(38), only IHC was used in this study to detect PD-L1 expression levels based on the recommendation in the NCCN Clinical Practice Guidelines(6), its ease of use, strong repeatability, and high accuracy(39, 40). Comparison among different detection methods should be considered in future research. In order to reduce sampling effect, only the area of the sample with more malignant cells, less differentiated cells, and less hemorrhage and necrosis was scored. The slides were scored blindly by 2 experienced pathologists to further improve the reliability of the PD-L1 expression levels. Third, the patient cohorts were heterogeneous in terms of PET/CT image acquisition. However, this can be viewed as a strength, as this heterogeneity decreases the possibility of overfitting to a particular subset of tumors or imaging parameters, and thus will result in a model that is more robust and transportable. Fourth, the stage distribution was different between the SPH and the HLM cohorts, as the HLM cohort was mostly advanced stage patients. However, the DLS was not correlated with tumor volume (*p=*0.10 for whole SPH cohort, *p=*0.79 for HLM-PD-L1-test cohort), and the DLS obtain high AUCs of 0.90(95%CI:0.85-0.97,p<0.001) among the subset of SPH patients with advanced stage, which suggest that stage doesn’t affect the final DLS prediction. Fifth, due to the small sample sizes of the prospective cohort, different HRs were observed for some clinical factors (BMI and ECOG). Though BMI was significant variable in univariable analysis, it was not significant in the multivariable Cox regression analysis for PFS and OS prediction. The C-index (0.88, 95%CI: 0.84-0.93) for the DCB prediction model with BMI was not statistically significantly different (p=0.09) compared to the DCB prediction model without BMI. As such, we opted to report a more parsimonious model where BMI was not included in the DCB, PFS and OS prediction model. From **Supplemental Tables S9** and **S10**, ECOG was included in the prediction models, but it didn’t affect the final C-indices of the prospective and external test cohorts based on the prediction results. Finally, more IO-treated patients with PD-L1 IHC score will be needed to improve the PD-L1 prediction model, and validate the efficiency of the DCB, PFS and OS prediction models.

In conclusion, an effective and stable deeply learned score to predict PD-L1 expression status was identified and may serve as a prognostic biomarker for immunotherapy response. Because images are routinely obtained and are not subject to sampling bias per se, we propose that the individualized risk assessment information provided by these analyses may be useful as a future clinical decision support tool pending in larger prospective trials.

## Materials & Methods

### Study population

In this multi-institutional study, five cohorts of patients were firstly accrued from two institutions: the Shanghai Pulmonary Hospital (SPH), Shanghai, China and H. Lee Moffitt Cancer Center & Research Institute (HLM), Tampa, Florida. The detailed inclusion criteria are provided in **Fig 1** and Supplemental S1. Among these, the SPH retrospective cohort, which was split into training (N=284) and validation (N=116) cohorts randomly by 70-30%, and the retrospective HLM cohort with PD-L1 status (N=85) were used for training and testing the DLS to predict PD-L1 expression; one IO-treated retrospective cohort (N=128) and one IO-treated prospective cohort (N=49) were used to investigate and validate the association of the DLS and clinical characteristics on the clinical outcomes. Additionally, a sixth cohort (N=35) from the third institution, James A. Haley Veterans’ Hospital (VA), Tampa, Florida, was curated as an external validation of the DLS and the prognostic models.

The progression of the distinct IO-treated cohorts used to investigate the association of the DLS and clinical characteristics with the clinical outcome including DCB (PFS>6month (41)), PFS, and OS, were defined using Response Evaluation Criteria in Solid Tumors (RECIST1.1)(42). The index date for both OS and PFS was the date of initiation of immunotherapy.

This study was approved by the Institutional Review Boards at SPH, University of South Florida (USF) and VA, and was conducted in accordance with ethical standards of the 1964 Helsinki Declaration and its later amendments. The requirement for informed consent was waived, as no PHI is reported.

### ^18^F-FDG PET/CT Imaging

Detailed acquisition parameters for the ^18^F-FDG PET/CT imaging for each cohort are presented in **Supplemental Table S11**. All PET images were converted into SUV units by normalizing the activity concentration to the dosage of ^18^F-FDG injected and the patient’s body weight after decay correction.

### PD-L1 expression by immunohistochemistry

The detailed information of IHC staining for PD-L1 expression is provided in Supplemental S2. For both SPH and HLM-PD-L1 cohort, the platform of Dako Link 48 and the antibody of Dako 22C3 were used for PD-L1 staining to quantify the presence of PD-L1. The level of PD-L1 expression was presented as a tumor proportion score (TPS), which is the percentage of viable tumor cells showing membrane PD-L1 staining relative to all viable tumor cells and is given as 0%, 1-49% and ≥50%, and PD-L1 positivity was defined as ≥1% of TPS(3, 43, 44).

### Development of the deeply learned score (DLS)

The pipeline for the PD-L1 expression prediction small-residual-convolutional-network (SResCNN) model is presented in **Supplemental Fig S4**. To train this model, the regions of interest (ROIs) of the PET and CT images from SPH were selected by experienced nuclear medicine radiologist (L.J) after registration using ITK-SNAP(45) on the condition that entire tumor and its peripheral region were included (**Supplemental Fig S5)**. To reduce the effect of the difference between the central slice and peripheral slices, only the ROIs with area larger than the 30% of the maximum area of this patient were regarded as valid ROI patches. The valid ROI patch was resized to 64×64 pixels by cubic spline interpolation, and constructed a three-channel hyper image together with their fusion image (alpha-blending fusion(46), α=1, **Supplemental Fig S6**). This hyper-image was input into the SResCNN model, and a deeply learned score (DLS) representing the PD-L1 positivity could be yielded after a sequential activation of convolution and pooling layers. To develop a robust prediction, the average DLSs of all valid slices including tumor tissue fed into the SResCNN model with equal weight was regarded as the final PD-L1 positive probability of the tumor. Details of the building, training, optimization and application methods were provided in **Supplementary S3**. The implementation of this model used the Keras toolkit and Python 3.5. The same pipeline (available at https://github.com/lungproject/lungio) was performed by an experienced radiologist (Y.S) on the three HLM cohorts and external VA cohort to obtain the DLS based on the guideline. Given there were minor differences between the different radiologists in selecting the ROIs, ROIs within the SPH-validation cohort were also selected by Y.S again to validate the reproducibility of DLS.

### Visualization of the SResCNN model

To further understand the prediction processing and explore the biological underpinnings of the deep learning feature, intermediate activation layers were firstly visualized to assess how the network carries the information from input to output(47). Additionally, the Gradient-weighted Class Activation Mapping (Grad-CAM) was used to understand the importance of each neuron for a decision of PD-L1 positive or negative, and produce a coarse localization map highlighting the important regions in the image for predicting the target concept (PD-L1 positive or PD-L1 negative) by using the gradient information of target concept flowing into the last convolutional layer of the SResCNN model. And the reconstructed maps were named as positive and negative filters later, which were also used to evaluate the class discrimination (28). Besides, unsupervised hierarchical clustering was performed on the deeply learned features (i.e., the output of global average pooling, N=256) to create a heatmap to show their distinguishable expression pattern among different patients. The clusters formed were based purely on the similarities and dissimilarities among the patients by the expressions of the deeply learned features.

### Statistical analysis

The Wilcoxon signed-rank test and Fisher’s exact test were used to test the differences for continuous variables and categorical variables, respectively. The area under the receiver operating characteristics curve (AUC), accuracy, sensitivity, specificity, and the 95% confidence interval (CI) by the Delong method(48) were used to assess the ability of DLS in discriminating between positive and negative PD-L1 expression. The median value of the DLS from the SPH-training patches was used as the cut-off. The inter-rater agreement of DLS estimations were calculated by intraclass correlation coefficient (ICC) between two radiologists. The correlation between different metadata (including age, BMI, Sex, stage, smoking status, ECOG, and SUVmax) and molecular biology (including histology, *EGFR, ALK, ROS1*, necrosis, and PD-L1 TPS) was analyzed by Spearman’s rank correlation coefficient. The details of necrosis quantification were shown in supplemental S4). Comparison of the magnitude of two correlations was performed with a software package named cocor(49).

In the IO-treated cohorts, the patients were clustered into high-DLS and low-DLS groups with the obtained cut-off, and survival analyses were performed using Kaplan-Meier method and Cox proportional hazards model. And further multivariable models, including the risk factors selected in univariate analysis according to the significance, were determined for the prediction of DCB, PFS and OS, which were evaluated using C-indices. Z test was applied to compare the differences between different models. To rigorously assess the quality of the study design, the radiomic quality score (RQS) was calculated(50) (Supplemental S5 and **Supplemental Table S12**). Two-sided p-values of less than 0.05 were regarded as significant and all statistical analyses were conducted with IBM SPSS Statistics 25 (Armonk, New York) and MATLAB R2019a (Natick, MA).

## Data Availability

The PET/CT imaging data and clinical information are not publicly available for patient privacy purposes, but are available from the corresponding authors upon reasonable request (R.J.G, and M.B.S).

## Acknowledgments

This study was funded by U.S. Public Health Service research grant U01 CA143062 and R01 CA190105 (principal investigator Dr. Gillies). This material includes work supported with resources and the use of facilities at the James A. Haley Veterans’ Hospital.

